# Long-term air pollution exposure and diabetes risk in American older adults: a national cohort study

**DOI:** 10.1101/2021.09.09.21263282

**Authors:** Maayan Yitshak Sade, Liuhua Shi, Elena Colicino, Heresh Amini, Joel D Schwartz, Qian Di, Robert O Wright

## Abstract

**Background:** Type 2 diabetes is a major public health concern. Studies show that both genetics and environmental exposures play a significant role in developing the disease. We assessed the association between air pollution and first documented diabetes occurrence in a national U.S. cohort of older adults to estimate diabetes risk.

**Methods:** We included all Medicare enrollees 65 years and older in the fee-for-service program, part A and part B, in the contiguous United States (2000-2016). Participants were followed annually until the first recorded diabetes diagnosis, end of enrollment, or death (264,869,458 person-years). We obtained annual estimates of fine particulate matter (PM_2.5_), nitrogen dioxide (NO_2_), and warm-months ozone (O_3_) exposures from highly spatiotemporally resolved prediction models. We assessed the simultaneous effects of the pollutants on diabetes risk using survival analyses. We repeated the models in cohorts restricted to ZIP codes with air pollution levels not exceeding the national ambient air quality standards (NAAQS) during the study period.

**Results:** We identified 10,024,879 diabetes cases of 41,780,637 people (3.8% of person-years). The hazard ratio (HR) for first diabetes occurrence was 1.074 (95% CI 1.058; 1.089) for 5 µg/m^3^ increase in PM_2.5,_ 1.055 (95% CI 1.050; 1.060) for 5 ppb increase in NO_2_, and 0.999 (95% CI 0.993; 1.004) for 5 ppb increase in O_3_. Both for NO_2_ and PM_2.5_ there was evidence of non-linear exposure-response curves with stronger associations at lower levels (NO_2_ ≤ 40 ppb, PM_2.5_ ≤ 9 µg/m^3^). Furthermore, associations remained in the restricted low-level cohorts. The O_3_-diabetes exposure-response relationship differed greatly between models and require further investigation.

**Conclusion:** Exposures to PM_2.5_ and NO_2_ are associated with increased diabetes risk, even when restricting the exposure to levels below the NAAQS set by the U.S. EPA.

## INTRODUCTION

Type 2 diabetes (T2D) is a major public health concern rising rapidly, with the number of people diagnosed with the disease worldwide more than doubling in the past 20 years ^1^. T2D may cause major complications, including blindness, cardiovascular damage, and premature mortality ^2^, it is therefore important to identify determinants of the disease.

Like many chronic conditions, type I diabetes (T1D) and T2D have a genetic component, but genetics alone explains only a small portion of the variance. Studies show that in T2D, which represents over 99% of diabetes cases globally ^3^, genetics and environmental exposures play a significant role ^4^. In the last few decades, environmental research focused primarily on behavioral factors, such as inactivity and diet. However, in recent years, cardiometabolic risk was linked to various environmental components, including the social environment ^5,6^, environmental pollutants ^3,7^, and, more specifically, air pollution ^8-13^. A recent meta-analysis concluded that each ten µg/m^3^ increase in long-term fine particulate matter (PM_2.5_) exposure was associated with an 8% increase in T2D prevalence risk, and each ten µg/m^3^ increase in long-term nitrogen dioxide (NO_2_) exposure was associated with a 7% increase in T2D prevalence. O_3_ was not included in that analysis since the number of studies was limited ^14^.

These positive associations were observed in other systematic reviews as well ^15,16^. However, since it is challenging to investigate the effects of the environment on chronic conditions, current research has many limitations. These limitations include potential bias in the outcome and exposure assessment, inability to separate incidence and prevalence cases, healthy survivor bias, and the inability to account for cardiometabolic risk factors and co-morbidities ^15^.

Our study addresses many of these limitations by investigating the association between particulate and gaseous air pollution and first documented diabetes. Our use of highly spatiotemporally resolved exposure models reduces bias due to exposure misclassification. In addition, unlike most national studies or chronic air pollution effects, we use comprehensive chronic conditions data on Medicare enrollees across the U.S. to reduce misclassification of diabetes and approximate incident cases.

In this study, we aim to assess the association between long-term air pollution exposure and first documented diabetes occurrence, focusing on the effect of air pollution exposure below the U.S. Environmental Protection Agency (EPA) national ambient air quality standards (NAAQS).

## METHODS

### Study population

We included all Medicare enrollees who were 65 years and older in the fee-for-service (FFS) program, part A (hospital insurance) and part B (medical insurance), in the contiguous U.S. between the years 2000-2016. We limited the data to person-years included in these programs because the algorithm used to identify chronic conditions utilizes claims covered by these three programs. We entered participants into the cohort on January 1 after they became Medicare participants and followed participants for each calendar year until the first recorded diabetes diagnosis, end of enrollment in either of the mentioned Medicare programs, or death – whichever came first. To avoid gaps in follow-up, once enrollment in the FFS, Medicare part A or B programs was terminated, those participants were no longer included in the cohort even if they renewed their enrollment in later years.

To better approximate incident cases, we excluded individuals diagnosed with diabetes before or in their first year of enrollment. We obtained information on diabetes status from the chronic conditions warehouse database. Diabetes was identified using an algorithm that incorporates claims indicating that an individual received a healthcare service for diabetes. The algorithm combines inpatient, outpatient, skilled nursing facilities, home health claims, or carrier claims (primarily doctor visits) ^17^. To be diagnosed with diabetes, beneficiaries must have at least one claim with the international classification of disease codes of diabetes (ICD-9 or ICD-10) from either inpatient, skilled nursing facility, home health, or part B services within two years.

This study was approved by the Centers for Medicare & Medicaid Services (CMS) under the data use agreement (#RSCH-2020-55733), the Institutional Review Board of Emory University (#STUDY00000316), and the Institutional Review Board of Mount Sinai (STUDY 20-01344), and a waiver of informed consent was granted. The Medicare dataset was stored and analyzed in the Rollins High-Performance Computing (HPC) Cluster at Emory University, in compliance with Health Insurance Portability and Accountability Act (HIPAA).

### Exposures

Our study was focused on three principal air pollutants linked to cardiometabolic health: PM_2.5_, O_3_, and NO_2_. PM_2.5_ are fine inhalable particles, smaller than 2.5 micrometers, comprising a mixture of solids and liquids. NO_2_ is an air pollutant that originates mostly from traffic and high-temperature combustion processes ^18^. O_3_ at the ground level is formed naturally and following a chemical reaction - where air pollutants emitted from sources such as traffic, industry, and wildfires (i.e., nitrogen oxides and volatile organic compounds) react with sunlight and organic gases, principally from vegetation ^19^.

We obtained predictions of PM_2.5_ (24-hour average, µg/m^3^), NO_2_ (daily 1-hour maximum, ppb) and ozone (daily maximum of 8-hour average, ppb) exposures from validated prediction models calibrated to measurements at approximately 2000 monitoring stations using an ensemble of three machine learners (neural network, random forest, and gradient boosting) that provided daily estimates for a 1 km^2^ grid of the contiguous U.S. ^20-22^. In brief, each machine learning algorithm incorporated more than 100 predictor variables from satellite data, land-use information, weather data, and chemical transport model simulations. The predictions of the three learners were then integrated using a generalized additive model-based geographically weighted-averaging technique. The model was calibrated using daily pollutants concentrations measured at EPA monitoring sites and demonstrated excellent model performance (average cross-validation R^2^=0.89, 0.84, and 0.86 for annual predictions of PM_2.5_, NO_2_, and O_3_, respectively). To align with the Medicare data, we aggregated gridded exposures to ZIP codes by averaging predictions of grid cells within each ZIP code annually. For ozone, we averaged the exposure only during the warmer months of each year (May – October), a commonly specified time window to examine associations with health outcomes ^23^. We selected this window because ozone formation is increased in warmer months due to its reaction with sunlight.

### Covariates

The Medicare data is a dynamic cohort that includes individual-level information on the participants’ sex, race, age, Medicaid eligibility, and date of death. We additionally obtained the following ZIP code level covariates from the U.S. Census: median household income, population density, percent home renters, percent of residents with no high school education, and percent of the population self-identified as black. We obtained data from available U.S. Census years and the annual American Community Survey, and extrapolated values for missing years. We aggregated annual summer, and winter temperature ZIP code means from gridded Daymet 1-km models ^24^.

### Statistical analysis

We investigated the effect of the three air pollutants simultaneously using a cox-equivalent re-parameterized Poisson survival approach ^25^. To allow for strata-specific baseline hazard functions, we stratified the models by follow-up year, calendar year, ZIP code, sex, race (white, black, other), age, and Medicaid insurance. We calculated diabetes counts in each follow-up year, calendar year, and ZIP code within strata specified by these individual characteristics, using the log of the corresponding total person-time as an offset. This approach is proven to be equivalent to a time-varying cox survival model using the Anderson-Gill formulation ^25^. We used the m-out-n bootstrap method to account for spatial autocorrelation of neighboring ZIP codes.

This methods samples ZIP codes randomly for each bootstrap replicate, and estimates statistically robust confidence intervals ^26^. First, we fit a multi-pollutant model using linear terms for the three pollutants. Then, to allow for nonlinear exposure-response curves, we used penalized spline functions for each of the pollutants. We fit three multipollutant models in which we used a penalized spline to estimate the exposure-response curve for one pollutant, and linear terms for the adjusted pollutants. We adjusted the models for annual summer and winter mean temperatures, population density, median household income, percent home renters, percent of residents without a high school diploma, and percent black population.

To assess the effect of air pollution concentrations below the NAAQS, we created three restricted low-exposure subsets comprised of individuals who were always exposed to exposure levels lower than the national standards within the study period (i.e., PM_2.5_ <12 μg/m^3^, NO_2_ <53 ppb). As there is no annual standard for O_3,_ we selected 50 ppb as a threshold to approximate the current World Health Organization global interim target 1 for peak-season average O_3_ concentration ^27^.

#### Sensitivity analyses

The exclusion of individuals who are not enrolled in the FFS, part A, and Part B programs can potentially induce a selection bias in the study. Biased results can also occur due to competing mortality risks. To avoid selection bias, we used inverse probability weights. Probabilities of enrollment in the cohort and probability of not dying were modeled, accounting for the subjects’ age, sex, race, Medicaid eligibility, and the ZIP code population density, percent population under the poverty line, percent population without a high school diploma, percent black and Hispanic population. We calculated these probabilities using pooled logistic regressions. We then calculated the weight by multiplying the inverse probability of enrolling in the three programs with the inverse probability of being alive and calculated the averaged weight within each stratum of follow up year, calendar year, ZIP code, and individual characteristics. We then repeated the models incorporating the weights and compared the exposure-response curves with and without weights.

## RESULTS

We have included 264,869,458 person-years of 41,780,637 people. The mean age was approximately 76 years, 60% were women, and 90% were white. We observed 10,024,879 diabetes cases (3.8% of person-years) (**Table 1**). The mean and IQR values of the air pollutants were as follows: PM_2.5_ 10.1 µg/m^3^ (4.2 µg/m^3^), O_3_ 43.2 ppb (7.0 ppb), and NO_2_ 18.9 ppb (13.7 ppb). The correlation between the exposures was low to moderate (**Supplementary Table 1**), with the highest correlation observed between PM_2.5_ and NO_2_ (r=0.44).

**Table 1.** Population characteristics (264,869,458 person years)

| Population characteristics | Summary statistics |
| --- | --- |
| **<u>Individual characteristics</u>** |  |
| Age, Mean (S.D.) | 75.97 (7.7) |
| Female sex, n (%) | 158,138,886 (59.7) |
| Race, n (%) |  |
| White | 238,995,206 (90.2) |
| Black | 14,986,036 (5.7) |
| Other | 10,888,216 (4.1) |
| Medicaid insurance, n (%) | 24,986,082 (9.4) |
| Diabetes, n (%) | 10,024,879 (3.8) |
| Hypertension, n (%) | 96,411,807 (36.4) |
| Ischemic heart disease, n (%) | 57,842,331 (21.8) |
| Death, n (%) | 1,125,608 (0.4) |
| **<u>ZIP code characteristics</u>** |  |
| Percent poverty, Mean (SD) | 0.1 (0.1) |
| Population density, Mean (SD) | 2,650.1 (7,854.1) |
| Median house value, Mean (SD) | 195,301.5 (159,465.5) |
| Percent black population, Mean (SD) | 0.1 (0.2) |
| Median household income, Mean (SD) | 53,618.0 (22,608.6) |
| Percent black population, Mean (SD) | 0.3 (0.1) |

Using linear terms, we observed an increased diabetes risk associated with 5 µg/m^3^ increase in PM_2.5_ (HR=1.074, 95% CI 1.058; 1.089) and 5 ppb increase in NO_2_ (HR=1.055, 95% CI 1.050; 1.060). No linear association was observed with O_3_ (HR 0.999,95% CI 0.993; 1.004). We then used penalized spline functions to allow for nonlinear exposure-response curves and found evidence of nonlinear associations with the three pollutants. Both for NO_2_ and PM_2.5_ we observed stronger associations at the lower ends of the pollutants’ distributions. For PM_2.5_ the associations were stronger for levels<8.2 µg/m^3^ (the 25^th^ percentile), followed by a plateaued association. For NO_2_ the associations were stronger for levels < 36 ppb (94^th^ percentile), followed by attenuated associations. The O_3_ exposure-response curve was highly nonlinear with mostly negative associations (**Figure 1**). The associations with NO_2_ and PM_2.5_ were similar in the single pollutant models and the multivariate model. The O_3_ exposure-response curve differed greatly between the single and multipollutant models. Unlike the negative associations seen in the multivariate model, there was no clear trend in the association with the pollutant in the single pollutant model (**Supplementary Figure 1**).

**Figure 1.**
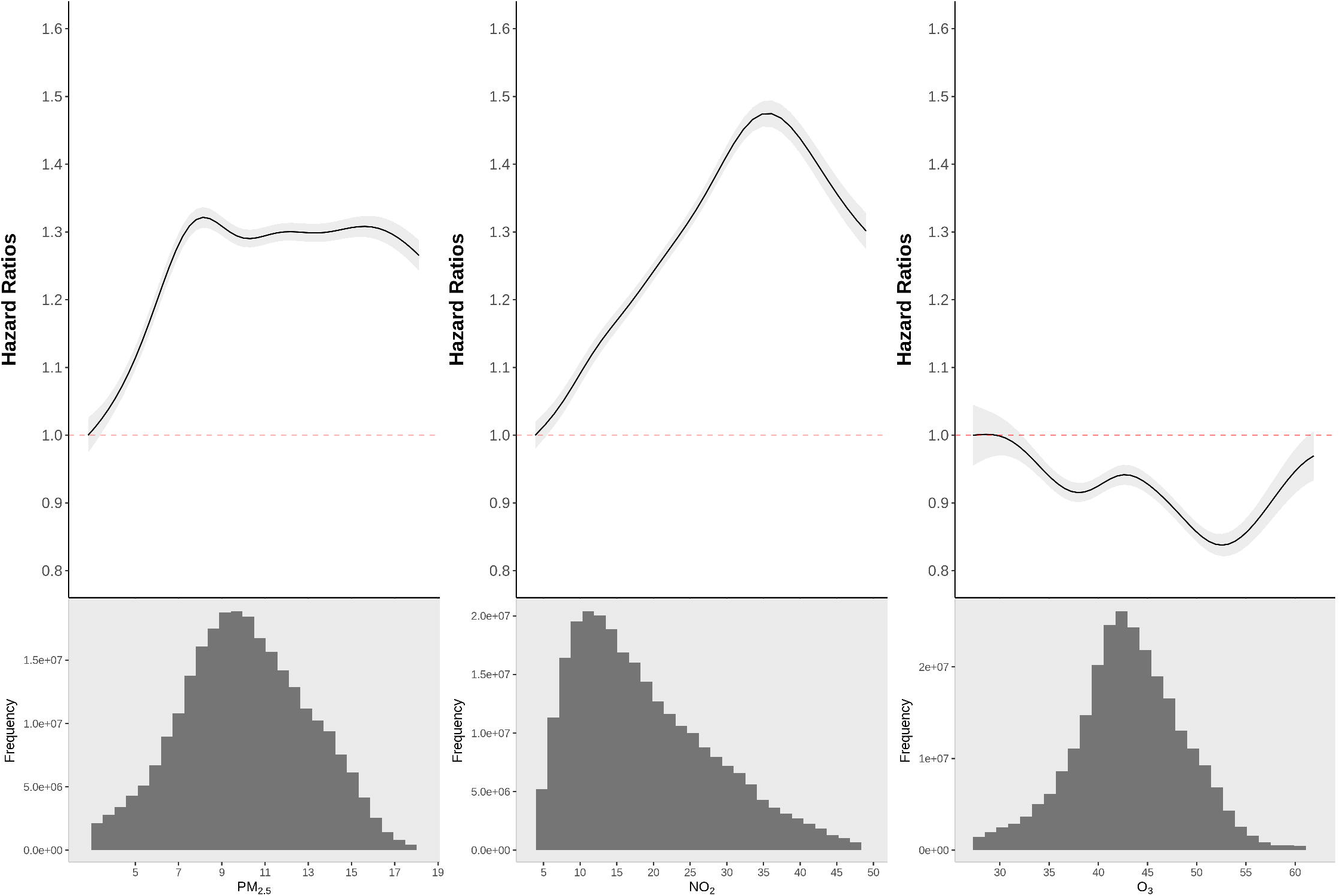
The association between PM_2.5_, NO_2_, and warm-months O_3_ exposure and diabetes occurrence. Figure 1 shows the exposure-response curves for the association between PM_2.5_, NO_2_, and warm-months O_3_ and diabetes occurrence. We show the curve from the 0.5^th^ percentile of PM_2.5_, i.e., with 0.5% poorly constrained extreme values excluded. Results are obtained from a multivariate model, adjusted for age, race, sex, Medicaid insurance, annual ZIP code means of summer and winter temperature, and annual ZIP code level sociodemographic variables. The model also includes a random intercept for each ZIP code and a spline function of year.

Of the analytical dataset, 33.7% (89,519,417 person-years), 96.4% (255,575,750 person-years), and 54.4% (144,225,079 person-years) were only exposed to low-level annual PM_2.5_, annual NO_2_, and warm-season O_3_ during the study period, respectively. High-pollution ZIP codes excluded from the restricted datasets were mostly located in the mid-west, southeast, and the mid-Atlantic U.S. regions. We estimated the nonlinear exposure-response curves for the three pollutants in multivariate regressions in these restricted cohorts, and the associations with PM_2.5_ and NO_2_ remained similar and significant. For O_3_, there was no clear association with diabetes and the direction of the associations differed across the distribution of the pollutant (**Figure 2**).

**Figure 2.**
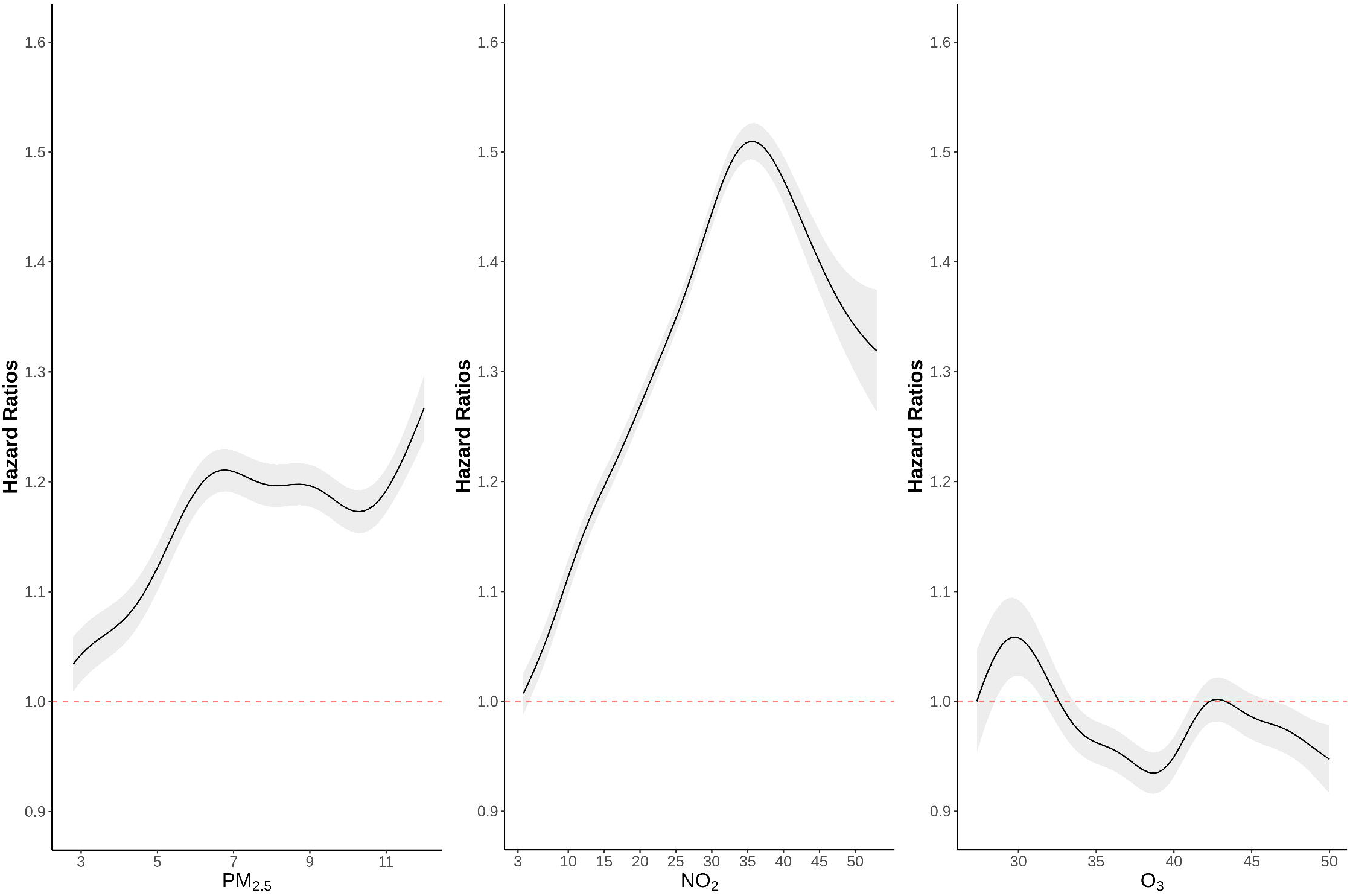
The association between low-levels air pollutants exposure and diabetes occurrence. Figure 2 shows the exposure-response curves for the association between PM_2.5_, NO_2_, and warm-months O_3_ and diabetes occurrence, in subsets, restricted to ZIP codes in which the annual exposures levels have not exceeded the national ambient air quality standards during the study period (i.e., PM_2.5_<12 µg/m^3^, NO_2_<35 ppb, or O_3_<50 ppb). We show the curves from the 0.5^th^ percentile of NO_2_, i.e., with 0.5% poorly constrained extreme values excluded. Results are obtained from a multivariate model, adjusted for age, race, sex, Medicaid insurance, annual ZIP code means of summer and winter temperature, and annual ZIP code level sociodemographic variables. The model also includes a random intercept for each ZIP code and a spline function of year.

**Supplementary Table 2** shows the baseline characteristics of those included and excluded from the analytical dataset. Person-years included in the analytical dataset were of older people. In addition, the proportion of whites and women was larger, and the proportion of death was much smaller. We conducted a sensitivity analysis and repeated the models including inverse probability weights accounting for the probability of being enrolled in the cohort and alive. The results of the two models were very similar (**Supplementary Figure 2)**.

## DISCUSSION

In a large-scale, national cohort, we observed higher risks for diabetes associated with increases in NO_2_ and PM_2.5_ exposures, even when restricting the data to exposure levels below the NAAQS set by the U.S. EPA. Results regarding the association with O_3_ were inconclusive.

T2D is characterized by high blood glucose levels and increased insulin resistance leading to vascular damage and metabolic dysfunctions ^29^. A growing body of literature links ambient air pollution exposure to diabetes risk, as concluded in many recent systematic reviews ^15,16^. Studies also found associations between air pollution exposure and markers of potential underlying pathways of this association: lower insulin sensitivity ^30^, and higher fasting glucose ^30,31^. Different mechanisms were proposed to explain the links between air pollution exposure and T2D. Oxidative stress is considered a major pathway to this association ^3^. PM exposure is associated with oxidative stress, which can lead to lipid peroxidation, reduction of antioxidants, and activation of pro-inflammatory processes ^9,32^. These, in turn, have a major role in the development and progression of metabolic syndrome and diabetes in particular ^33,34^. Another suggested pathway is PM-induced mitochondrial dysfunction, which decreases brown adipose tissue ^35^. Since stimulated brown adipose tissue regulates glucose homeostasis and insulin resistance using glucose and free fatty acids during thermogenesis ^36,37^, this may systematically interfere with glucose metabolism and insulin sensitivity. Studies also found indications of oxidative stress and mitochondrial disfunction following NO_2_ and O_3_ exposure ^38-41^.

Our study shows evidence of increased diabetes risk associated with long-term PM_2.5_ and NO_2_ exposures. Like our findings, a 2019 meta-analysis concluded that the odds for T2D were 1.03 and 1.05 times higher for incremental increases of 10 µg/m^3^ in PM_2.5_ and NO_2_ exposures, respectively ^16^. Studies assessing the association between O_3_ and diabetes are much more limited, and the direction of the observed associations is inconsistent and varies greatly depending on the model covariates ^42^. For example, a longitudinal cohort of 13,548 individuals in China found a 5.7% increase in diabetes incidence hazard associated with a ten µg/m^3^ increase in annual O_3_ exposure ^43^. However, two recent long-term studies did not find a significantly increased diabetes risk associated with O_3_ exposure in multipollutant models accounting for particulate matter ^44,45^ and NO_2_ ^44^ exposures. Moreover, a 2021 review concluded that evidence on the association between O_3_ exposure and diabetes is insufficient to infer causality ^42^. Our study also showed inconclusive results, possibly related to residual confounding, exposure measurement error, or the complexities of simultaneously estimating the effects of multiple air pollutants.

The Clean Air Act was last amended in 1990 and requires the U.S. EPA to set NAAQS that mitigate any harmful consequences of air pollution to human health and the environment ^28^. However, recent studies suggest that these thresholds are insufficient to protect human health. There is evidence of increased mortality risk associated with air pollution levels below the NAAQS ^46-51^. In some analyses, the associations observed at the lower exposure distribution range were stronger than the higher range of the exposure distribution ^50,51^. This may be attributed to larger measurement error at the rarer higher exposure concentrations that is likely to attenuate the overall effect observed when considering the full ranges of the pollutant exposures in the analysis ^52^. Regarding PM_2.5_, it is also possible that the composition of the particles is different for low and high pollution days ^46^.

We limited our study sample to ZIP codes with lower air pollution levels to simulate a scenario where air pollution levels never exceeded thresholds considered safe by the U.S. EPA. The major findings of our study are the nonlinear associations and the harmful air pollution effects observed even from levels below the NAAQS set by the EPA. These results suggest that NO_2_, and PM_2.5_ exposures below the NAAQS are associated with increased cardiometabolic risk. Like our findings, Paul et al. ^53^ also observed associations between moderate exposure concentrations of PM_2.5_ and NO_2_ and diabetes mortality and incidence risks. Moreover, they report greater PM_2.5_ effects on diabetes incidence risk in lower pollution concentrations.

Our study has several limitations. First, there are differences between people included and excluded from the cohort. This is a limitation of all studies that analyze claims data of the Medicare cohort. However, our sensitivity analysis suggests that our results were not biased due to differential probabilities of enrollment or death. Second, the Medicare data does not provide information on subtypes of diabetes or individual confounders such as BMI, smoking, physical activity, or lifestyle. However, since exposure is assigned on a ZIP code level, neighborhood factors are more likely to confound the associations in our study. These neighborhood factors were accounted for in our models. Finally, we might have had exposure misclassification errors like other air pollution studies. However, the use of highly spatiotemporally resolved exposure models reduces this error.

In conclusion, assessing the simultaneous effects of particulate and gaseous air pollutants in a national cohort, we found increased diabetes risk associated with PM_2.5_ and NO_2_ exposure. The observed effects remained when restricting the data to exposure levels below the NAAQS. For ozone, the effects were inconclusive and require further investigation. Since current studies of the link between air pollution and diabetes are scarce and often limited in quality or sample size, this national study may add robust evidence important for inferring the causal link between air pollution exposure and the development of diabetes.

## Data Availability

The Medicare data includes protected health information and therefore cannot be shared. Researchers who wish to use the data may contact the center for Medicare and Medicaid services directly.

## ACKNOWLEDGEMENTS

This study was supported by the HERCULES Center (P30 ES019776), the Mount Sinai transdisciplinary center on early environmental exposures (P30 ES023515), the National Institute on Aging (NIA/NIH R01 AG074357), the National Institute of Environmental Health Sciences (R21 ES032606, R01 ES032242, 5U2CES026555-03, and R01 ES013744, P30 ES000002, R01 ES032418), and the United States Environmental Protection Agency (US EPA) (RD-83587201). Its contents are solely the responsibility of the grantee and do not necessarily represent the official views of the US EPA. Furthermore, the US EPA does not endorse the purchase of any commercial products or services mentioned in the publication. Finally, H.A. is supported by Novo Nordisk Foundation Challenge Programme: Harnessing the Power of Big Data to Address the Societal Challenge of Aging (NNF17OC0027812).

