## Supplemental material for "Long-term air pollution exposure and diabetes risk in American older adults: a national cohort study"

**Supplementary Table 1.** The correlation between the exposures and climate variables.

| | $PM_{2.5}$ | $NO_2$ | Ozone | Summer temperature |
|---|---|---|---|---|
| $NO_2$ | 0.44 | | | |
| Ozone | 0.27 | 0.18 | | |
| Summer temperature | 0.1 | -0.04 | 0.23 | |
| Winter temperature | -0.04 | -0.04 | -0.07 | 0.62 |

**Supplementary Table 2.** Baseline characteristics of person years by status of enrollment in the fee for service, part A, and part B programs.

| | Included (n=432,826,576 – 61%) | Excluded (n=283,734,324 - 39%) |
|---|---|---|
| Age, Mean (Q1; Q3) | 76 (70; 82) | 74 (67; 80) |
| Female sex, n (%) | 254,446,733 (58.7%) | 156990829 (55.3%) |
| Race, % (n) | | |
| White | 381,308,542 (88.0%) | 230,692,691 (81.3%) |
| Black | 29,667,751 (6.8%) | 26,666,991 (9.3%) |
| Other | 19,535,981 (4.5%) | 23,060,733 (8.1%) |
| Missing | 2,314,302 (0.7%) | 3,313,909 (1.3%) |
| Medicaid eligible, % (n) | 50792506 (11.7%) | 37,428,498 (13.1%) |
| Death, % (n) | 2141710 (0.4%) | 28664286 (10%) |

**Supplementary Figure 1. The associations between $PM_{2.5}$, $NO_2$, $O_3$ and diabetes: results of single pollutant models.**

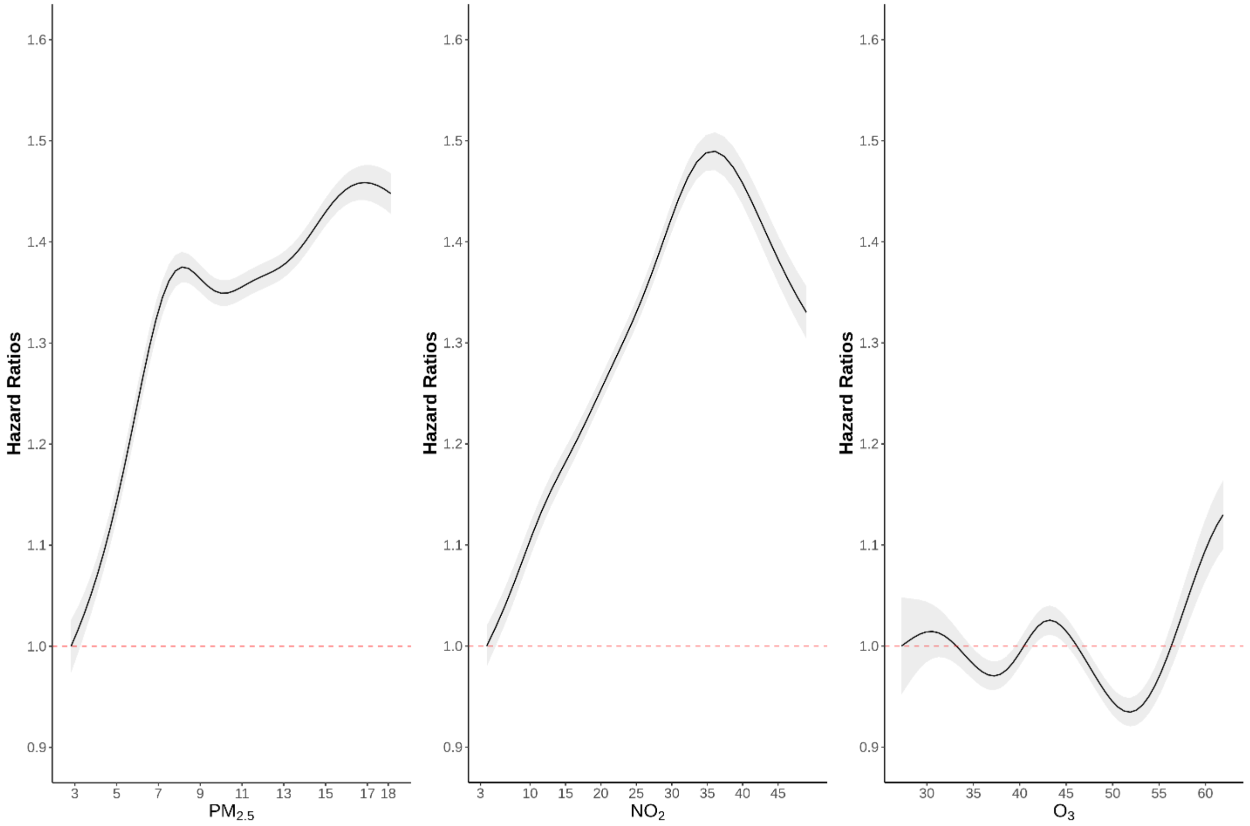


This figure shows the exposure-response curves for the associations with $PM_{2.5}$, $NO_2$, and warm-months $O_3$, from 0.5$^{th}$ percentile-99.5$^{th}$ percentile of pollutants, i.e., with 1% poorly constrained extreme values excluded. Results are obtained from three single-pollutant models, adjusted for age, race, sex, Medicaid insurance, annual ZIP code means of summer and winter temperature, and annual ZIP code level sociodemographic variables. The model also includes a random intercept for each ZIP code and a spline function of year.

**Supplementary Figure 2.** Comparison of results – the main analysis (red line) versus a sensitivity analysis weighted by the inverse probability of being in the cohort (blue line).

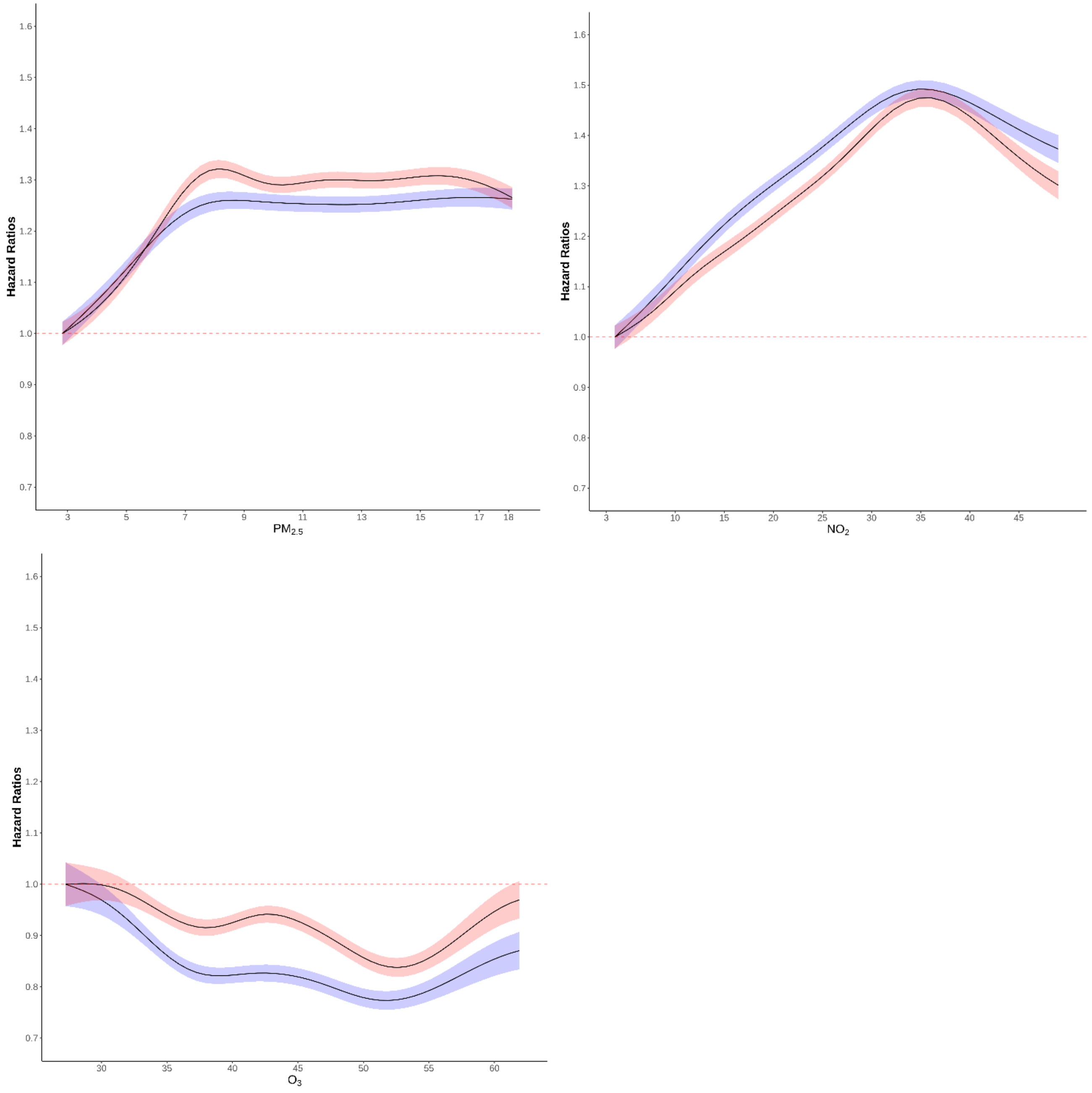


This figure shows the exposure-response curves for the associations with $PM_{2.5}$, $NO_2$, and warm-months $O_3$, from 0.5$^{th}$ percentile-99.5$^{th}$ percentile of pollutants, i.e., with 1% poorly constrained extreme values excluded. Results are obtained from a multivariate model, adjusted for age, race, sex, Medicaid insurance, annual ZIP code means of summer and winter temperature, and annual ZIP code level sociodemographic variables. The model also includes a random intercept for each ZIP code and a spline function of year.
